# Development and Validation of a Pan-*Plasmodium* Molecular Assay as a Confirmatory Method for Malaria Blood Donor Screening

**DOI:** 10.64898/2025.12.22.25342854

**Authors:** Pamela Milani, Li Wen, Leilani Montalvo, Sonia Bakkour Coco, Clara Di Germanio, Vanessa Bres, Manisha Yadav, Kristin Livezey, José Eduardo Levi, Jeffrey Linnen, Michael P. Busch

## Abstract

**Background:** Transfusion-transmitted malaria remains a concern in non-endemic regions due to asymptomatic parasitemia in donors with prior residence in or travel to malaria-endemic areas. Nucleic acid testing (NAT)–based screening has been proposed to mitigate this risk, and supplemental assays are needed to confirm reactive donations. We developed a pan-*Plasmodium* reverse transcription quantitative PCR (RT-qPCR) assay with species identification capability as a supplemental tool for malaria donor screening.

**Study design and methods:** The assay targets a conserved region of the 18S rRNA shared by all five human-infecting *Plasmodium* species and is compatible with whole blood lysed using Grifols’ Parasite Transport Medium (PTM). Analytical performance was evaluated using *in vitro* transcripts and infected red blood cells (iRBCs), with the limit of detection (LoD) determined by probit analysis. Specificity was assessed against *Babesia microti* and 300 non-exposed U.S. donor samples. Clinical sensitivity was evaluated using infected specimens; species identification was performed by sequencing of RT-qPCR amplicons.

**Results:** The assay demonstrated high amplification efficiency (97.2%) and linearity (R² = 0.99). The 95% LoD was 5.3 iRBCs/mL (95% CI: 3.2–8.6), comparable to the Procleix Plasmodium Assay. Clinical sensitivity was 100% across all five *Plasmodium* species, and clinical specificity was 100% (95% CI: 99–100%), with no cross-reactivity with *Babesia*. Amplicon sequencing enabled accurate species-level identification of all sequenced specimens.

**Discussion:** This assay provides a sensitive confirmatory tool for malaria NAT-based donor screening. Its compatibility with PTM lysates and species identification capabilities supports regulatory applications and research into asymptomatic parasitemia in semi-immune donors.

## INTRODUCTION

Transfusion-transmitted malaria (TTM) remains a risk in non-endemic regions, as donors with prior residence in or travel to malaria-endemic areas may harbor asymptomatic parasitemia^1–5^. Unlike vector-borne malaria, in which sporozoites first undergo hepatic replication, TTM results from the direct introduction of parasitized red blood cells into the bloodstream, enabling rapid progression to severe disease, particularly in immunologically naïve or immunocompromised recipients^3^.

In the United States (U.S.) and Europe, approximately 20 TTM cases have been reported over the past two decades^4,6^. Current U.S. donor deferral policies mandate a 3-month deferral for travelers to malaria-endemic regions and a 3-year deferral for former residents of endemic areas or individuals with a history of malaria. Although these measures substantially reduce risk, they rely on accurate donor recall of travel history and prior malaria exposure and may not identify all donors who are asymptomatically infected, particularly among prior residents of endemic regions. Retrospective analyses of TTM cases in the U.S. indicate that most reported transmissions occurred in donors who were eligible at the time of donation, having exceeded required deferral intervals, and were associated primarily with chronic, low-density parasitemia capable of persisting asymptomatically for years or even decades following exposure, particularly after childhood infection or repeated exposure in endemic settings^4^.

At the same time, questionnaire-based deferral significantly impacts the blood supply. Approximately 150,000 donors are deferred annually in the U.S. due to malaria-related criteria, many of whom pose minimal actual risk^7,8^. This over-deferral reduces blood availability and may disproportionately exclude donors from genetically diverse populations. Given these factors, laboratory-based screening has been explored as a malaria risk mitigation strategy. However, conventional methods are inadequate: thick blood smears and antigen-based assays lack the sensitivity to detect low-density parasitemia, while serologic testing—although more sensitive—cannot distinguish past from active infection because antibodies may persist after parasite clearance, leading to unnecessary deferrals of non-infectious donors.

To overcome these limitations, regulatory agencies have increasingly supported nucleic acid test (NAT)–based screening strategies for malaria. In 2022, the Procleix Plasmodium Assay^9^ (Grifols Diagnostic Solutions Inc.) for donor screening received the CE mark certification. In 2024, the U.S. Food and Drug Administration (FDA) approved the cobas Malaria test (Roche Molecular Systems). This was followed in January 2025 by FDA draft guidance recommending selective NAT as an alternative or adjunct to questionnaire-based deferrals^10^. A key advancement in molecular approaches is the use of ribosomal RNA (rRNA) as a target. With thousands of rRNA copies per parasite, compared to fewer than 10 genomic DNA copies^11,12^, rRNA-based assays offer enhanced sensitivity, particularly for detecting low-level parasitemia in asymptomatic individuals. Both the cobas Malaria test and the Procleix Plasmodium Assay leverage this principle.

As these platforms progress through regulatory approval and implementation, validated supplemental confirmatory methods become essential for supporting licensure studies, confirming reactive screening results, and informing donor counseling. Numerous reverse-transcription qPCR (RT-qPCR) assays targeting *Plasmodium* 18S rRNA have been developed for clinical diagnosis, epidemiological surveillance, and controlled human malaria infection studies^12–16^. These assays typically report limits of detection of approximately 20 parasites/mL^12,15^. By comparison, the cobas Malaria test and the Procleix Plasmodium Assay report analytical sensitivities in the range of 2–6 parasites/mL, depending on the *Plasmodium* species^9^. While improved sensitivity of RT-qPCR assays has been demonstrated by processing large blood volumes^17^, they have not been directly validated against donor-screening platforms under matched experimental conditions to assess sensitivity comparability, nor have they been evaluated to directly operate on the chemically stabilized whole-blood lysates used in donor screening NAT workflows.

To address this gap, we developed a pan-*Plasmodium* RT-qPCR assay as a confirmatory tool to support regulatory licensure studies of the Procleix Plasmodium Assay. The assay is compatible with whole-blood lysates prepared in Grifols Parasite Transport Medium (PTM), which lyses erythrocyte membranes to release parasites while preserving and stabilizing RNA. It targets a conserved region of the 18S rRNA transcript shared by all five human-infecting *Plasmodium* species, uses oligonucleotide sequences orthogonal to those of the Procleix assay, and matches its input volume to enable direct analytical comparison. Importantly, the RT-qPCR amplicon contains species-specific polymorphisms that permit species identification by post-amplification sequencing, providing a practical tool for epidemiologic investigation and donor counseling. In addition, assay performance is monitored using amplification of an endogenous human RNA target as an extraction control, enabling assessment of sample quality and supporting Cq-based, semi-quantitative interpretation of *Plasmodium* signal levels relative to the endogenous control.

We aimed to evaluate the analytical and clinical performance of this RT-qPCR assay and assess its suitability as a confirmatory method supporting malaria NAT implementation in blood donor screening.

## MATERIALS AND METHODS

### Study design

Analytical performance—including amplification efficiency, linearity, and limit of detection—was evaluated using contrived dilution panels prepared from flow cytometry-quantified *P. falciparum*-infected red blood cells and synthetic RNA transcripts. Analytical specificity was evaluated both in silico and experimentally by testing nucleic acids from *Babesia microti*—including RNA from a murine infection model and a high-copy synthetic DNA standard—to confirm the absence of cross-reactivity with this transfusion-relevant intraerythrocytic parasite. Clinical performance was assessed using whole-blood specimens from healthy donors and from individuals naturally infected with *P. falciparum*, *P. vivax*, *P. ovale*, or *P. malariae*. The assay’s ability to detect all five human-infecting *Plasmodium* species was further evaluated using *P. knowlesi*-infected macaque red blood cells. Species discrimination capability was assessed through sequencing of RT-qPCR amplicons.

### Sample collection

Non-infected blood samples, obtained from Creative Testing Solutions (Tempe, AZ) and Vitalant (Scottsdale, AZ), were collected with written, voluntary consent for research use as part of the standard blood donation process (Advarra protocol Pro00030878). Whole-blood samples from individuals naturally infected with *P. falciparum*, *P. vivax*, *P. malariae*, and *P. ovale* were obtained from the Wadsworth Center Parasitology Laboratory as de-identified residual diagnostic samples under IRB exemption category 4 (45 CFR 46.104 (d)(4)) for secondary research use (New York State Department of Health IRB protocol). No additional informed consent was required for either sample source.

Ring-stage *P. falciparum* culture was procured from New York Blood Center Enterprises (Rye, NY).

*P. knowlesi*-infected red blood cells were obtained from macaques housed and cared for at the University of Georgia, an AAALAC-accredited and PHS-certified institution. All blood collection procedures were reviewed and approved by the University of Georgia Institutional Animal Care and Use Committee (IACUC). Research involving the *Babesia microti* model was performed under approval and oversight of the IACUC at PMI Preclinical, LLC, under Animal Welfare Assurance A3367-01.

### Sample preparation and RNA extraction

EDTA-anticoagulated whole blood (0.9 mL) was mixed with 2.7 mL of Procleix PTM. For RNA extraction, 500 µL of lysate was processed using TRI Reagent (ThermoFisher Scientific) with 1-bromo-3-chloropropane phase separation, followed by isopropanol precipitation and ethanol washes. RNA was resuspended in 100 µL of nuclease-free water.

### Preparation of *in vitro* transcripts

*P. falciparum in vitro* transcripts (IVTs) containing the 18S rRNA target region were generated by cloning a synthetic gBlocks fragment (Integrated DNA Technologies) into pcDNA3.1 and transcribing with the mMESSAGE mMACHINE Kit (ThermoFisher Scientific). IVT concentration was quantified by spectrophotometry, and copy number was calculated based on transcript length and molecular weight.

IVTs for *P. vivax*, *P. ovale*, *P. malariae*, and *P. knowlesi* were provided by Grifols Diagnostic Solutions.

### Pan-*Plasmodium* RT-qPCR assay

The assay was designed to ensure that primers and probe sequences exhibit 100% identity across all five human-infecting *Plasmodium* species, while containing mismatches with *Babesia* species, as confirmed by BLAST analysis. The primer and probe sequences were as follows:

Pl-rRNA-1037F: 5’-TCAAGAACGAAAGTTAAGGGAGT-3’

Pl-rRNA-1075-Probe: 5’-/56-FAM/CCGTCGTAA/ZEN/TCTTAACCATAAACTAT/3IABkFQ/-3’

Pl-rRNA-1202R: 5’-CCAAAGACTTTGATTTCTCAT-3’

We also included amplification of an endogenous control (Ribosomal protein lateral stalk subunit P0 [RPLP0]) for monitoring extraction efficiency. The primer and probe sequences were as follows:

RPLP0-Fw: 5’-GCTTCCTGGAGGGTGTCC-3’

RPLP0-Probe: 5’-/56-FAM/TGCCAGTGT/ZEN/CTGTCTGCAGATTGG/3IABkFQ/-3’

RPLP0-Rv: 5’-GGACTCGTTTGTACCCGTTG-3’

*Plasmodium* and RPLP0 assays were run in separate reactions. Primer and probe concentrations and ratios were optimized to achieve high amplification efficiency. RT-qPCR was performed using the SuperScript III Platinum One-Step qRT-PCR Kit (ThermoFisher Scientific). Each 25 μL reaction contained final concentrations of 1X PCR buffer, 1.0 μM each of forward and reverse primers, 0.15 μM probe (for the *Plasmodium* assay) or 0.25 μM probe (for the RPLP0 assay), 1X SuperScript enzyme mix, and 10 μL of RNA template. Amplification was performed on a LightCycler 480 Instrument II (Roche Diagnostics) using the following thermal profile: reverse transcription at 50°C for 30 minutes, followed by initial denaturation at 95°C for 2 minutes, and then 45 cycles of 95°C for 15 seconds and 60°C for 1 minute.

### Quality controls and acceptance criteria

Each run included positive process controls (PTM-lysed iRBCs at 100 and 50 iRBCs/mL), negative process controls (non-infected whole-blood lysates), positive template controls (IVTs at 5,000 and 500 copies/µL), and no-template controls. Valid runs required non-reactive negative controls, expected positive control performance, and an RPLP0 Cq less than 22. This threshold was derived from RNA extraction reproducibility studies (typical RPLP0 Cq 18–20 cycles) with an added conservative safety margin.

### *Babesia microti* 18S rRNA qPCR and RT-PCR assay

Total RNA was extracted from 200 µL of whole-blood samples collected from the *Babesia* murine infection model. RNA was eluted in 100 µL. Reverse transcription was performed using *Babesia*-specific primers and MuLV Reverse Transcriptase (Applied Biosystems). qPCR was performed using either the resulting cDNA or *Babesia* DNA standards (Quantitative Genomic DNA from *Babesia* microti strain Gray, ATCC) as template. All qPCR reactions were performed using SYBR Green I according to the protocol outlined in Bloch et al., 2013^18^.

### Species identification

RT-qPCR amplicons were purified, quantified, and subjected to bidirectional Sanger sequencing using the same forward and reverse primers used for RT-qPCR amplification. Consensus sequences were queried against the NCBI nucleotide database using BLASTN (megablast algorithm), with results filtered to retain only human-infecting *Plasmodium* species. For each sample, the highest-scoring human-validated BLAST hits were recorded, and the corresponding species identification was extracted.

### Statistics

Statistical analyses were performed using R (version 4.4.3; R Foundation for Statistical Computing), with probit regression implemented using the *glm* function and the *dose.p* function from the MASS package.

## RESULTS

### RNA extraction reproducibility

We first assessed the reproducibility of the manual TRI Reagent-based RNA extraction protocol from PTM-lysed whole blood. We conducted 22 independent extractions using non-infected whole-blood lysates, plus two aliquots of *P. falciparum*-infected erythrocytes diluted to 100 and 50 iRBCs/mL. Each 500 µL aliquot underwent the complete extraction protocol followed by RT-qPCR amplification of the endogenous control gene RPLP0.

The RPLP0 Cq values demonstrated high reproducibility, with a mean of 18.45 (95% CI: 18.28–18.62), a coefficient of variation of 2.2%, and a range of 17.92–19.89. The Cq values of infected samples were not significantly different from uninfected samples (18.58 for 100 iRBCs/mL; 18.51 for 50 iRBCs/mL). This coefficient of variation is consistent with published assays^12^ and supports the reliability of downstream *Plasmodium* detection.

### Amplification efficiency and linearity

Amplification efficiency and linearity were first evaluated using serial dilutions of *P. falciparum* IVTs in water, independent of matrix effects. IVTs were tested using a six-point, ten-fold dilution series ranging from 5 × 10^8^ to 5 × 10^3^ copies/mL across eight independent runs. The assay showed high linearity (R^2^ = 0.99) and an amplification efficiency of 97.2% (Figure 1A). At the lowest concentration (5 × 10^3^ copies/mL), amplification remained detectable with a mean Cq of 36.72.

**Figure 1.**
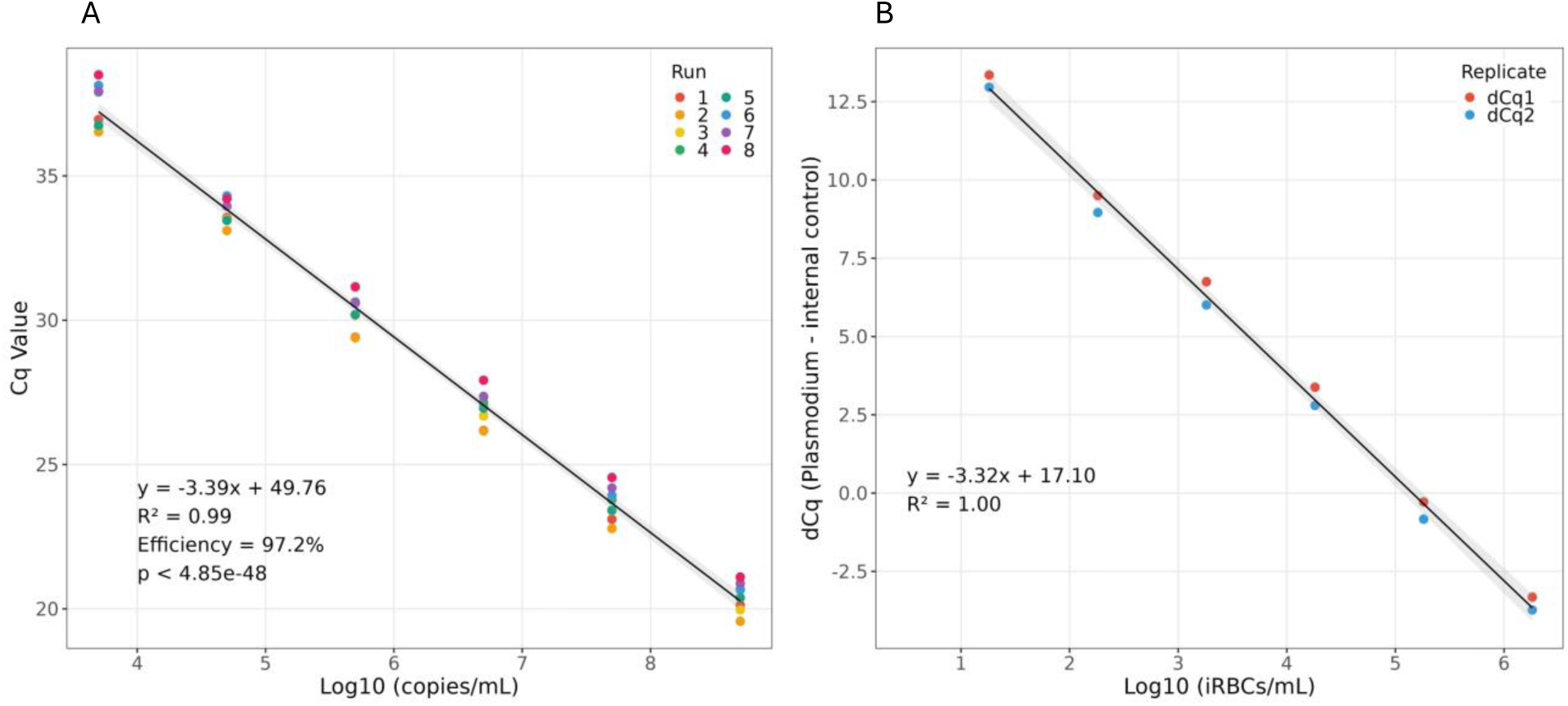
Linearity and amplification efficiency of the pan-*Plasmodium* RT-qPCR assay using *P. falciparum in vitro* transcripts (IVTs) and infected red blood cells (iRBCs). Standard curves were generated using serial dilutions of *P. falciparum* targets. (A) IVTs in water were tested across eight independent runs (colored dots, runs 1-8). (B) Serial dilutions of iRBCs in ABO-matched whole blood were tested in duplicate, with normalized Cq values (dCq = Cq Plasmodium - Cq endogenous control) plotted against concentration. Regression lines show linearity with statistical parameters displayed for each assay.

To evaluate performance under physiologically relevant conditions, we tested serial dilutions of ring-stage *P. falciparum* iRBCs in ABO-matched whole blood lysed with PTM buffer, ranging from 1.8 × 10^6^ to 18 iRBCs/mL. Cq values were normalized to RPLP0, and delta Cq (dCq) values were plotted against concentration. Regression analysis yielded an R^2^ of 1.00 and a slope of −3.32, indicating excellent linearity across the parasite dilution range in the relevant biological matrix (Figure 1B).

### Limit of detection

The limit of detection (LoD) was determined in accordance with the CLSI EP17-A2 guidelines^19^. An LoD panel was constructed by diluting flow cytometry-quantified ring-stage *P. falciparum* iRBCs into group-matched whole blood at six concentrations: 100, 30, 10, 3, 1, and 0.3 iRBCs/mL. At each concentration, 30 independent aliquots were lysed with PTM and tested by RT-qPCR. In parallel, 30 matched aliquots were tested using the Procleix Plasmodium Assay.

Probit regression was applied to model detection probability. For the RT-qPCR assay, the 50% LoD was 1.3 iRBCs/mL (95% CI: 1.0–1.6) and the 95% LoD was 5.3 iRBCs/mL (95% CI: 3.2–8.6). The Procleix assay demonstrated a 50% LoD of 0.4 iRBCs/mL (95% CI: 0.3–0.6) and a 95% LoD of 2.4 iRBCs/mL (95% CI: 1.3–4.3). Although point estimates for both LoD metrics were lower for the Procleix assay, overlap of the 95% LoD confidence intervals indicates comparable analytical sensitivity within this concentration range (Figure 2).

**Figure 2.**
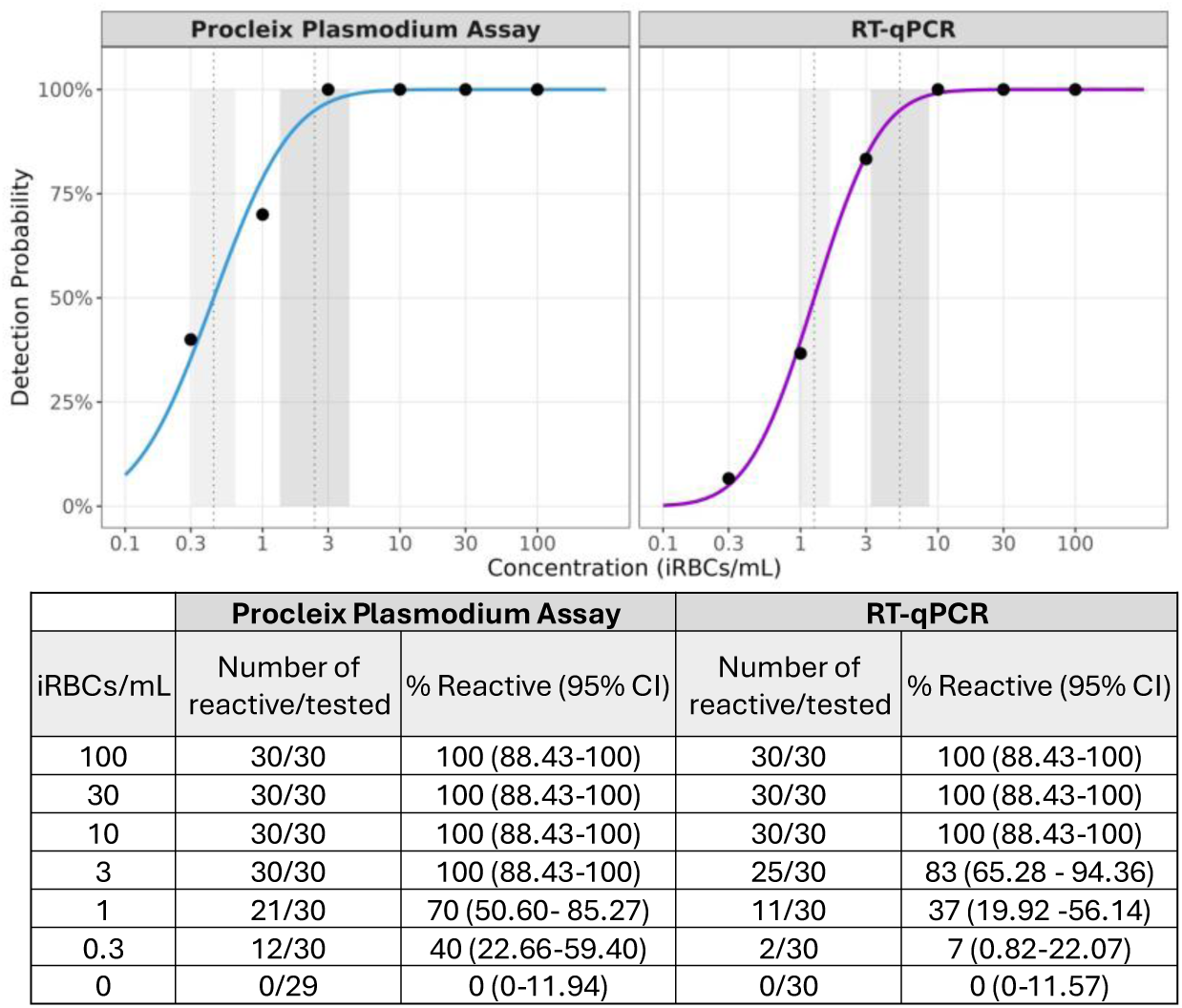
Limit of detection analysis for the Procleix Plasmodium Assay and the pan-*Plasmodium* RT-qPCR using probit regression. Detection probability curves were fitted using probit regression for both assays. Observed detection rates are shown as black dots at each iRBCs/mL concentration. The 50% and 95% limits of detection are indicated by vertical dotted lines, with shaded areas representing their 95% confidence intervals (CIs). The table in the bottom panel summarizes the number of reactive samples over the total tested, along with detection percentages and 95% CIs, at each concentration level for both assays.

### Analytical and clinical specificity

In silico alignment revealed sequence mismatches between the *Plasmodium*-specific primers and probe of our assay and the homologous regions in *Babesia* genomes. Nevertheless, we also confirmed analytical specificity experimentally. Total RNA was extracted from whole blood collected from a mouse model infected with *Babesia microti*. Additionally, a high-copy *Babesia microti* synthetic DNA template (4 × 10^4^ copies/µL) was included. Both samples were tested in duplicate using the pan-*Plasmodium* RT-qPCR assay. No amplification was observed in either *Babesia* sample, whereas a *Plasmodium* IVT control (5 × 10^4^ copies/µL) showed an average Cq of 25.33, as expected. Validity of the extraction and template material was confirmed using a separate qPCR assay with *Babesia*-specific primers. The *Babesia* DNA standard amplified with an average Cq of 20.13, and the mouse model-derived RNA yielded a Cq of 12.75 when using the *Babesia* molecular assay. These results experimentally confirmed that the pan-*Plasmodium* RT-qPCR assay does not cross-react with *Babesia* nucleic acids.

Clinical specificity was assessed using 300 human whole-blood PTM-lysate samples from healthy U.S. donors who were not deferred based on malaria residence or travel history. No false-positive *Plasmodium* signals were detected, yielding a clinical specificity of 100% (95% Clopper-Pearson CI: 99–100%). To ensure that non-reactive results were not attributable to inefficient RNA extraction, all samples were verified to have RPLP0 Cq values below the established acceptance threshold of 22 cycles, confirming adequate RNA recovery and assay validity.

### Detection of all human-infecting *Plasmodium* species

The ability of the RT-qPCR assay to detect all five *Plasmodium* species known to infect humans was evaluated using both synthetic material and clinical specimens. Initial testing was performed using IVTs for each species, spiked into PTM-lysed non-infected whole blood at three concentrations: 1,500, 3,000, and 6,000 copies/mL. Each IVT-spiked lysate underwent three independent RNA extractions, with three amplification replicates per extraction. The assay successfully detected all five species at all tested concentrations (Figure 3).

**Figure 3.**
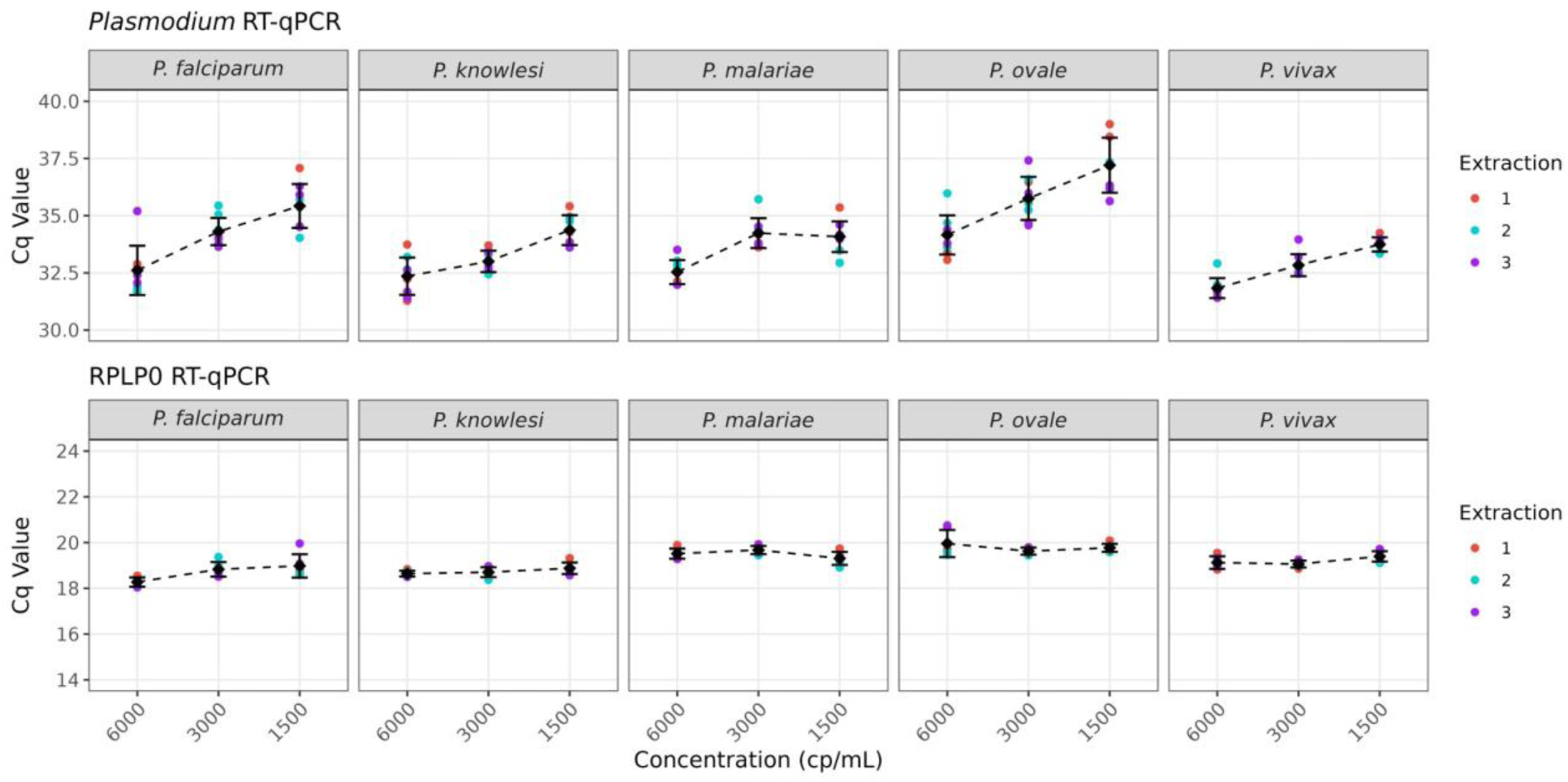
Detection of All Five *Plasmodium* Species in IVT-Spiked Whole-Blood Lysate. RT-qPCR detected all five *Plasmodium* species in IVT-spiked whole blood at 1,500, 3,000 and 6,000 copies/mL. Each condition was tested across three extractions and three amplification replicates. The assay showed consistent performance across all species and concentrations.

To further assess species detection under clinically relevant conditions, the assay was tested on 25 *Plasmodium*-positive whole-blood lysates representing all five species. The majority of these were clinical samples from individuals naturally infected with *P. falciparum*, *P. vivax*, *P. malariae*, or *P. ovale*, with species identity confirmed by the Wadsworth Center. The single specimen of *P. knowlesi*, consisting of infected macaque RBCs, had been confirmed positive via microscopy. Each lysate underwent three RNA extractions, followed by a single RT-qPCR amplification. The assay was reactive for all the samples, yielding a clinical sensitivity of 100% (95% Clopper-Pearson CI: 86.3–100%). Cq values ranged from 15.9 to 25.9 (Figure 4, top panel). In both the IVT spike-in and clinical sample testing, the internal control gene RPLP0 was consistently amplified with Cq values below the assay threshold (Figures 3 and 4, bottom panel).

**Figure 4.**
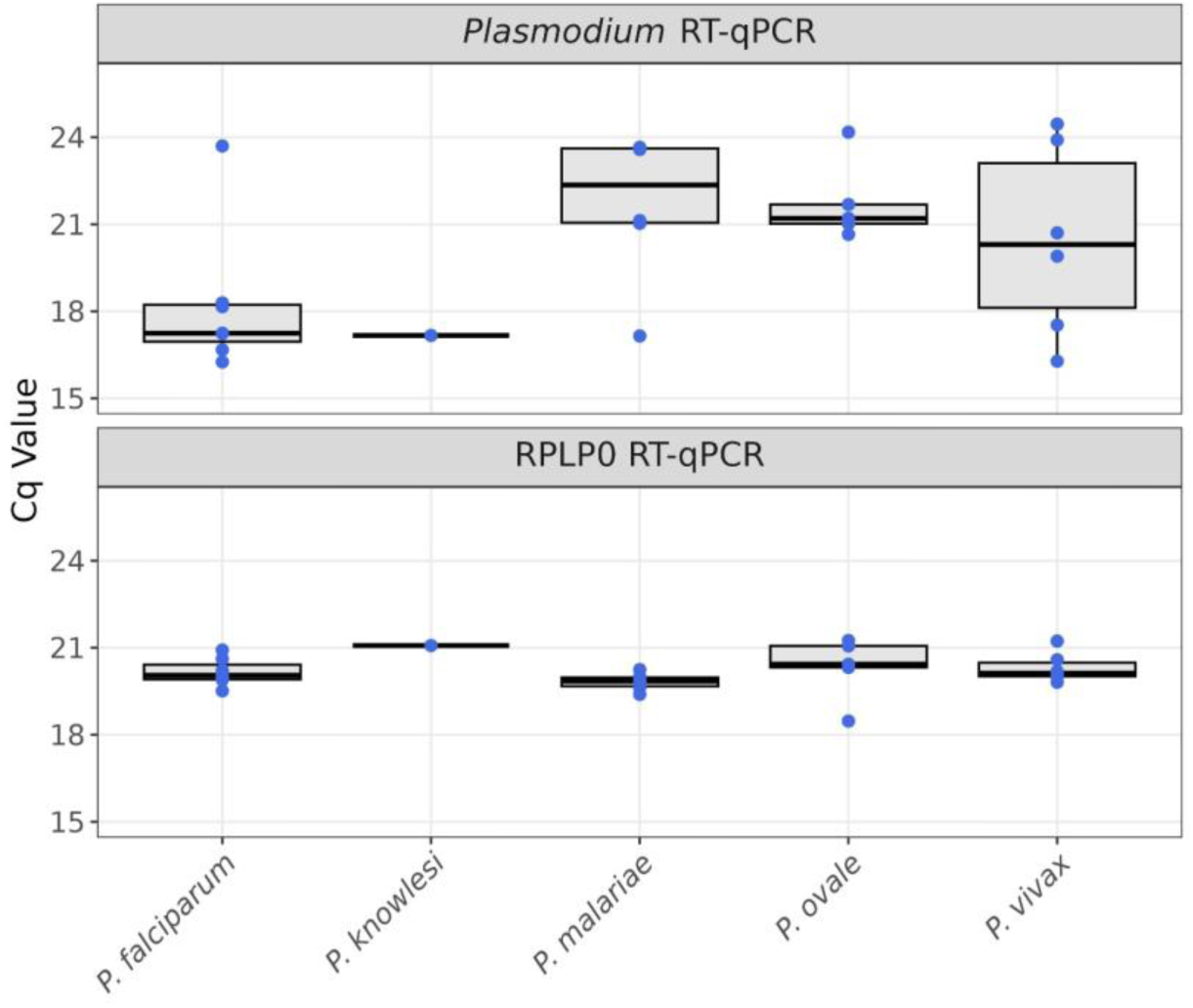
Clinical Sensitivity of the RT-qPCR Assay across *Plasmodium* Species. Box plots showing Cq values from RT-qPCR targeting *Plasmodium* RNA and the human RPLP0 control. Whole-blood lysates from 25 *Plasmodium*-positive specimens—covering all five human-infecting species—were extracted in triplicate, and Cq values shown represent the mean per specimen. The assay achieved 100% detection across all species.

### Species identification by amplicon sequencing

The primers and probe of the pan-*Plasmodium* RT-qPCR assay target highly conserved sites shared across all five human-infecting *Plasmodium* species; however, the internal amplicon region contains species-specific sequence mismatches (Figure 5A). This design enables post-amplification species identification through direct sequencing (Figure 5B). Following RT-qPCR, amplicons were purified (Figure 5C), subjected to bidirectional Sanger sequencing, and queried against the NCBI nucleotide database to assign species identity. To evaluate the accuracy of this approach, we analyzed 21 *Plasmodium*-positive clinical samples encompassing all five species. The sequencing-based identification method demonstrated 100% concordance with the reference laboratory results (Table 1).

**Figure 5.**
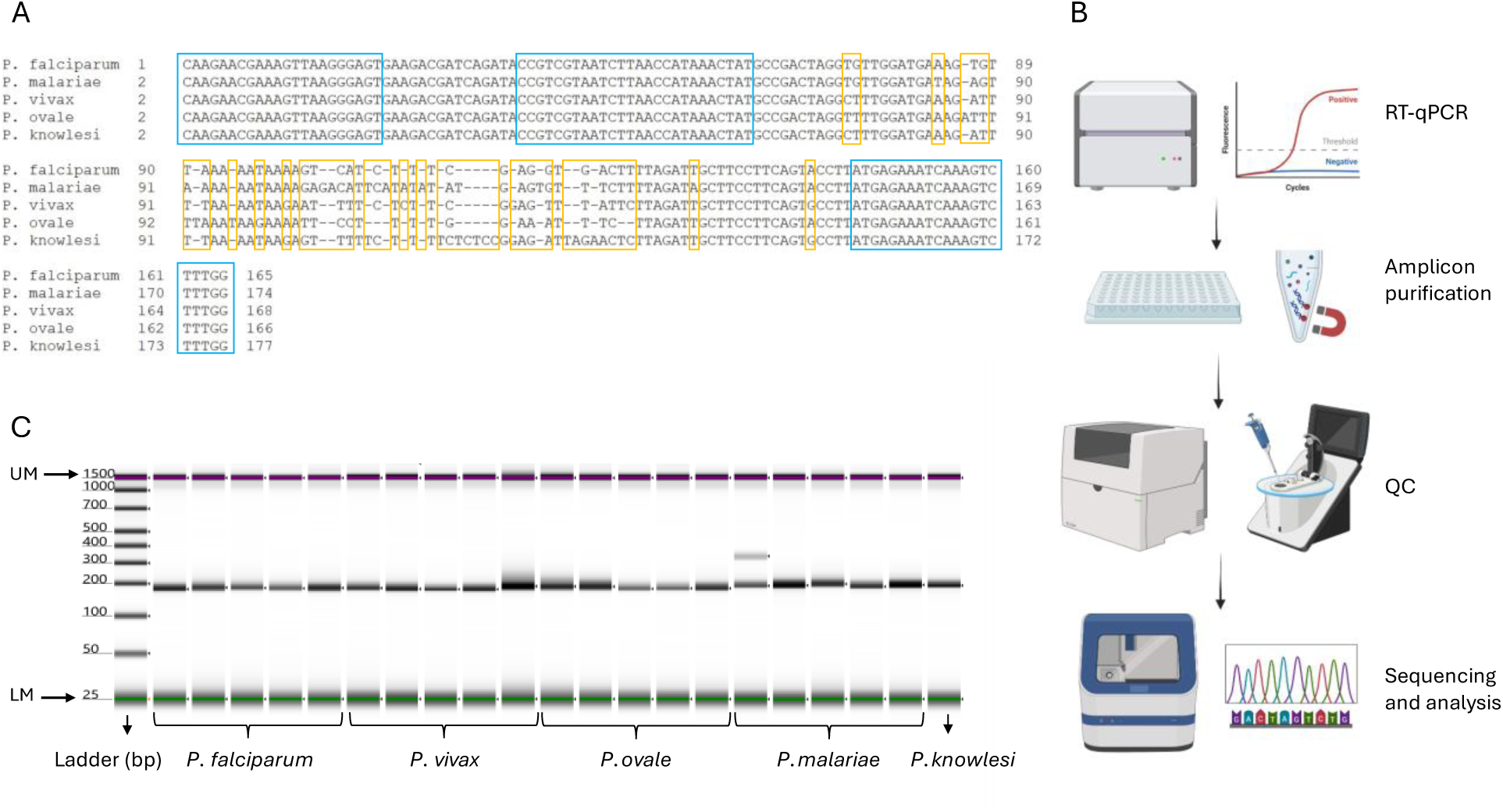
Species-Level Identification of *Plasmodium* via Post-Amplification Sequencing. (A) Sequence alignment of the RT-qPCR target region highlights conserved primer/probe binding sites (blue rectangles) across the five human-infecting *Plasmodium* species, while the internal amplicon region contains species-specific polymorphisms (orange rectangles). (B) Workflow schematics depicting the post-amplification process, including amplicon purification, quality control (QC) with the TapeStation System and NanoDrop, and bidirectional Sanger sequencing for species identification. This schematic was created using BioRender (BioRender.com). (C) TapeStation analysis of purified RT-qPCR products shows a single, distinct band for all samples, consistent with the expected ∼170 bp amplicon and confirming amplification specificity. One *P. malariae* sample displayed an additional nonspecific band of ∼300 bp; however, Sanger sequencing still produced accurate species identification for this sample. The 1,500 bp band corresponds to the upper marker (UM), and the 25 bp band corresponds to the lower marker (LM) of the TapeStation assay; both markers are indicated by arrows.

**Table 1.** Concordance between Species Identification by the Reference Assay and Amplicon Sequencing. *Plasmodium*-positive clinical samples (n=21) underwent bidirectional sequencing following amplification with our pan-*Plasmodium* RT-qPCR assay. Species-level identification was performed using BLAST analysis of consensus sequences. Forward and reverse reads consistently yielded concordant species identifications; for clarity of presentation, only the alignment with the highest percent identity is shown in the table. All 21 samples were correctly identified, resulting in 100% concordance with the species determinations from the reference laboratory.

|  |  | Alignment output of pan-*Plasmodium* RT-qPCR amplicon sequencing |  |  |
| --- | --- | --- | --- | --- |
| SampleID | Species identified by the reference assay | Definition | Identity (%) | Accession |
| LYSPFALSAMPLE1 | *P. falciparum* | Plasmodium falciparum isolate R73<br>18S ribosomal RNA gene, partial sequence | 99.09 | HQ283215 |
| LYSPFALSAMPLE2 | *P. falciparum* | Plasmodium falciparum isolate R73<br>18S ribosomal RNA gene, partial sequence | 95.06 | HQ283215 |
| LYSPFALSAMPLE3 | *P. falciparum* | Plasmodium falciparum isolate R73<br>18S ribosomal RNA gene, partial sequence | 100 | HQ283215 |
| LYSPFALSAMPLE4 | *P. falciparum* | Plasmodium falciparum isolate R73<br>18S ribosomal RNA gene, partial<br>sequence | 97.2 | HQ283215 |
| LYSPFALSAMPLE5 | *P. falciparum* | Plasmodium falciparum isolate R73<br>18S ribosomal RNA gene, partial<br>sequence | 99.16 | HQ283215 |
| LYSPKNOSAMPLE1 | *P. knowlesi* | Plasmodium knowlesi isolate<br>SABAH89.2 18S ribosomal RNA gene,<br>partial sequence | 96.74 | KT852895 |
| LYSPMALSAMPLE1 | *P. malariae* | Plasmodium malariae strain VEN02<br>18S ribosomal RNA gene, partial<br>sequence | 89.74 | KJ619945 |
| LYSPMALSAMPLE2 | *P. malariae* | Plasmodium malariae strain VEN02<br>18S ribosomal RNA gene, partial<br>sequence | 100 | KJ619945 |
| LYSPMALSAMPLE3 | *P. malariae* | Plasmodium malariae strain VEN02<br>18S ribosomal RNA gene, partial<br>sequence | 92.06 | KJ619945 |
| LYSPMALSAMPLE4 | *P. malariae* | Plasmodium malariae strain KOY13<br>18S ribosomal RNA gene, partial<br>sequence | 97.58 | KJ619942 |
| LYSPMALSAMPLE5 | *P. malariae* | Plasmodium malariae strain VEN02<br>18S ribosomal RNA gene, partial<br>sequence | 100 | KJ619945 |
| LYSPOVASAMPLE1 | *P. ovale* | Plasmodium ovale curtisi clone DC-1<br>18S ribosomal RNA gene, complete<br>sequence | 96.72 | KF696369 |
| LYSPOVASAMPLE2 | *P. ovale* | Plasmodium ovale curtisi clone DC-1<br>18S ribosomal RNA gene, complete<br>sequence | 95.35 | KF696369 |
| LYSPOVASAMPLE3 | *P. ovale* | Plasmodium ovale wallikeri clone GC-1<br>18S ribosomal RNA gene, complete<br>sequence | 100 | KF696359 |
| LYSPOVASAMPLE4 | *P. ovale* | Plasmodium ovale wallikeri clone GC-1<br>18S ribosomal RNA gene, complete<br>sequence | 100 | KF696359 |
| LYSPOVASAMPLE5 | *P. ovale* | Plasmodium ovale wallikeri clone GC-1<br>18S ribosomal RNA gene, complete<br>sequence | 100 | KF696359 |
| LYSPVIVSAMPLE1 | *P. vivax* | Plasmodium vivax isolate H15 18S<br>ribosomal RNA gene, partial sequence | 98.31 | HQ283224 |
| LYSPVIVSAMPLE2 | *P. vivax* | Plasmodium vivax isolate SV1 18S<br>ribosomal RNA gene, partial sequence | 100 | JQ627153 |
| LYSPVIVSAMPLE3 | *P. vivax* | Plasmodium vivax isolate SV1 18S<br>ribosomal RNA gene, partial sequence | 100 | JQ627153 |
| LYSPVIVSAMPLE4 | *P. vivax* | Plasmodium vivax isolate SV1 18S<br>ribosomal RNA gene, partial sequence | 98.17 | JQ627153 |
| LYSPVIVSAMPLE5 | *P. vivax* | Plasmodium vivax isolate H15 18S<br>ribosomal RNA gene, partial sequence | 93.02 | HQ283224 |

## DISCUSSION

The pan-*Plasmodium* RT-qPCR assay described here demonstrated excellent analytical performance, including high amplification efficiency and linearity across a broad dynamic range, and robust detection at low parasite densities. The assay produced no false positives among healthy donor samples, detected all five human-infecting *Plasmodium* species with complete sensitivity, and showed no cross-reactivity with *Babesia microti*—a key requirement given the transfusion relevance of this related pathogen, which is endemic in many regions of the U.S.

Although previously published RT-qPCR methods have proven effective for their intended clinical and research applications, direct comparison with donor-screening NAT platforms has been limited by differences in sample matrices and input volumes. The assay described here was purpose-built to address these limitations, operating on the same PTM-lysed specimens and matching the volume of the Procleix Plasmodium Assay to support direct, side-by-side assessment.

When tested using the same panels, the analytical sensitivity of the pan-*Plasmodium* RT-qPCR assay closely approximated that of the Procleix platform, with overlapping confidence intervals for the 95% limit of detection. This degree of sensitivity alignment is critical for confirmatory testing, minimizing the risk of discrepancies attributable to sensitivity differences. Operationally, compatibility with the Procleix specimen matrix eliminates the need for separate sample collection, streamlining logistics and ensuring that orthogonal testing is performed on material with preserved RNA integrity.

Beyond its role as a supplemental assay in regulatory settings, the pan-*Plasmodium* RT-qPCR assay has broader utility in malaria research, particularly for investigating infection dynamics and parasite persistence in semi-immune individuals. This study demonstrates that sequencing of RT-qPCR amplicons enables accurate discrimination of all five *Plasmodium* species. Species-level identification is especially relevant in this context, as *Plasmodium* species differ in persistence, relapse potential, and clearance kinetics^20–22^. Moreover, because clinical management and follow-up vary by species, sequence-based identification provides a practical and valuable tool to assist clinicians evaluating reactive donors^23^.

A quantitative version of this assay is under development for longitudinal monitoring of parasitemia in asymptomatic individuals. Notably, the performance of the current RT-qPCR assay—including its linear response and correlation of Cq values with input parasite levels in IVT and iRBC dilution panels—demonstrates the feasibility of deriving semi-quantitative estimates of parasite signal using external standard curves. The quantitative assay under development will build on this foundation by incorporating normalization to reference genes, a validated reportable range, and calibrated standards to enable precise and reproducible quantification across study batches. By detecting low-level fluctuations in serial samples, the assay is expected to support characterization of variability and clearance patterns in chronic infections. In addition, it may provide insights into the persistence of subclinical reservoirs and inform future studies on transmission risk and donor screening policies, including evaluation of the relationship between parasite levels in whole blood and detectable parasite sequences in plasma, the sample type used for malaria screening in Brazil^24^.

A current limitation of the assay is that both RNA extraction and RT-qPCR are performed manually. While operationally acceptable for orthogonal confirmation of a limited number of samples, this workflow may constrain scalability for larger research or screening efforts. Planned adaptation to automated liquid-handling systems is expected to reduce hands-on time and enable higher throughput.

In summary, these findings demonstrate the assay’s suitability for confirmatory testing and outline practical steps to extend its application to translational research.

## Data Availability

All data produced in the present study are available upon reasonable request to the authors

## ACKNOWLEDGMENTS

We thank Chester J. Joyner from the University of Georgia for providing the *Plasmodium knowlesi* sample.

